# Relationship Between Depression, Prefrontal Creatine and Gray Matter Volume

**DOI:** 10.1101/2021.02.16.21251840

**Authors:** Paul Faulkner, Susanna Lucini Paioni, Petya Kozhuharova, Natasza Orlov, David J. Lythgoe, Yusuf Daniju, Elenor Morgenroth, Holly Barker, Paul Allen

## Abstract

**Background:** Depression and low mood are leading contributors to disability worldwide. Research indicates that clinical depression may be associated with low creatine concentrations in the brain and low prefrontal gray matter volume. Because sub-clinical depression also contributes to difficulties in day-to-day life, understanding the neural mechanisms of depressive symptoms in all individuals, even at a sub-clinical level, may aid public health.

**Methods:** Eighty-four young adult participants completed the Depression, Anxiety and Stress Scale (DASS) to quantify severity of depression, anxiety and stress, and underwent ^1^H-Magnetic Resonance Spectroscopy of the medial prefrontal cortex and structural MRI to determine whole-brain gray matter volume.

**Results/Outcomes:** DASS depression scores were negatively associated with A) concentrations of creatine (but not other metabolites) in the prefrontal cortex, and B) with gray matter volume in the right superior medial frontal gyrus. Medial prefrontal creatine concentrations and right superior medial frontal gray matter volume were positively correlated. DASS anxiety and DASS stress scores were not related to prefrontal metabolite concentrations or whole-brain gray matter volume.

**Conclusions/Interpretations:** This study provides preliminary evidence from a representative group of individuals who exhibit a range of depression levels, that prefrontal creatine and gray matter volume are negatively associated with depression. While future research is needed to fully understand this relationship, these results provide support for previous findings which indicate that increasing creatine concentrations in the prefrontal cortex may improve mood and wellbeing.

**Declaration of Interest/Funding:** This research was partly funded by a British Academy/Leverhulme Trust Research Grant, awarded to PA.

## Introduction

Depression and low mood are leading contributors to disability worldwide, and can affect more than 300 million people at any one time (Britton, 2017). While most of the negative health and social effects of low mood are attributed to Major Depressive Disorder (MDD), individuals who experience sub-clinical depression (i.e. those who score below clinical thresholds on depression scales) also experience a significant impact on their daily functioning (Cuijpers et al., 2004). Importantly, the prevalence of sub-clinical depression/low mood may be increasing in society, and may now affect a higher percentage of society (24%) than clinical depression (Van Zoonen et al., 2015). Because depression may be best understood as a continuum of symptoms, and because sub-clinical depression may be one of the best predictors of future MDD (Rodriguez et al., 2012, Van Zoonen et al., 2015), understanding the mechanisms of sub-clinical depressive symptoms in non-clinical groups may contribute to improvements in public mental health.

To aid development of novel therapies for depression, studies have attempted to determine the neurochemical mechanisms of depressive symptoms, yet many have ignored the potential importance of brain creatine. This is partly because when using the primary method for quantifying this endogenous compound *in vivo* (^1^H-Magnetic Resonance Spectroscopy; ^1^H-MRS), research groups have often referenced their metabolite of interest to creatine to ‘correct’ for concentrations of other metabolites (e.g. Kumar et al., 2002), on the basis that creatine was considered to be stable within regions of interest and across individuals (Li et al., 2003). However, prefrontal creatine is influenced by cigarette smoking (Durazzo et al., 2016; Faulkner et al., 2020a), cocaine use (Chang et al., 1997), cannabis use (Prescot et al., 2011), anxiety (Yue et al., 2012) and schizophrenia (Öngür et al., 2009). Moreover, the relationship between brain creatine and depression is poorly understood. Specifically, no difference was found in anterior cingulate creatine metabolite concentrations between 19 depressed individuals diagnosed with MDD and 30 aged-matched controls (Auer et al., 2000), or in thalamic creatine concentrations between 18 young depressed individuals and 18 young non-depressed individuals (Mirza et al., 2005). Conversely, Kondo et al (2016) reported a weak yet significant relationship between depression scores and concentrations of creatine in the frontal cortex of 22 adolescent participants diagnosed with MDD (*p* = 0.030; actual effect size not reported). Because daily administration of 20g creatine monohydrate for four weeks can increase total brain creatine (see Allen (2012) for a review), and because creatine supplements can alleviate depressive symptomatology when taken daily for >2 weeks (Lyoo et al., 2012; Nemets and Levine, 2013), it is important to determine whether there is a relationship between prefrontal creatine concentrations and depression.

Structural neuroimaging studies have also indicated that depression and low mood are related to lower gray matter volume in the prefrontal cortex. Two meta-analyses of 20+ studies that examined brain structure using voxel-based morphometry (VBM) revealed that compared to healthy controls, depressed patients exhibit lower gray matter volume bilaterally in the medial prefrontal cortex and anterior cingulate cortex (Bora et al., 2012; Lai., 2013). However, it is currently unknown whether this depression-related low prefrontal gray matter volume is related to alterations in prefrontal creatine concentrations.

In the current study we examined the relationship between depression levels in a non-clinical sample and both prefrontal creatine metabolite concentrations (quantified using ^1^H-MRS) and gray matter volume (quantified using VBM). It was hypothesized that there would be a significant negative association between depression levels and both creatine concentrations and gray matter volume in the prefrontal cortex. For completeness, and on the basis of findings by Hasler et al (2007) and Auer et al (2000), who report that depressed individuals exhibit low prefrontal concentrations of glutamate and GABA metabolites, we also performed exploratory analyses to determine the relationship between depression levels and concentrations of all metabolites quantified by the ^1^H-MRS sequence.

## Methods

We report an analysis of data collected from two separate studies, both of which had the aim of collecting health-related MRI data in young adults. Both study protocols used the same MRI sequences for volumetric and ^1^H-MRS data (see below), and all data were acquired on the same 3T MRI scanner at the Combined Universities Brain Imaging Centre.

### Participants

Across both studies, 84 participants were recruited via print and online advertisements. Thirty-eight of these subjects participated in study 1, and 46 subjects participated in study 2. All participants gave written informed consent after receiving a detailed explanation of their study procedures (approved by the University of Roehampton Research Ethics committee), and were screened for eligibility. Exclusion criteria for both studies were: self-report of psychiatric diagnoses; current drug use/abuse or dependence (other than tobacco and cannabis use); history of neurological injury or disease, pregnancy and contraindications for MRI (e.g. metal implants). Data from study 1 were collected from November 2015 - March 2018, while data from study 2 were collected from September 2017 - December 2019.

### Questionnaire Measures

Participants completed a demographics form (developed in-house) to determine age, gender, level of education, self-reported psychiatric comorbidity, neurological disorder and use of tobacco, cannabis and illicit drugs. Exposure to cigarettes was inferred from the average number of cigarettes smoked per day as in previous research (e.g. Durazzo et al., 2016; Faulkner et al., 2018; 2019; 2020b). Participants also completed the Depression, Anxiety and Stress Scale (DASS), a 42-item questionnaire designed to quantify levels of depression, anxiety and stress; each of these emotional symptoms is assessed by summing the answers to fourteen 4-point Likert scale questions (Lovibond and Lovibond, 1995). On the depression subscale, a score of 0-9 denotes no depression, 10-13 mild depression, 14-20 moderate depression, 21-27 severe depression, while 28+ denotes extremely severe depression. On the anxiety subscale, a score of 0-7 denotes no anxiety, 8-9 mild anxiety, 10-14 moderate anxiety, 15-19 severe anxiety and 20+ denotes extremely severe anxiety. On the stress subscale, a score of 0-14 denotes no stress, 15-18 mild stress, 19-25 moderate stress, 26-33 severe stress and 34+ denotes extremely severe stress.

To determine the influence of age, gender, cigarette smoking and cannabis use on depression, anxiety and stress, an ANOVA was constructed, in which scores from the DASS depression, anxiety or stress subscales were added as the dependent variable (as relevant), and age, gender, the average number of cigarettes smoked per day, and the average number of cannabis joints smoked per day were all added as separate factors.

### ^1^H-MRS Data Acquisition, Pre-processing and Analysis

All ^1^H-MRS scans were acquired using the same 3T Siemens Magnetom TIM Trio MRI system using a 32-channel head coil. ^1^H-MRS *in vivo* spectra were acquired from the same 20 × 20 x 20mm voxel located in the right medial prefrontal cortex (typical location shown in *Figure 1A*). The structure and function of the medial prefrontal cortex are related to the clinical features of depression (e.g. Farb et al., 2011); this voxel placement therefore allowed us to test our own hypotheses pertaining to relationships of depression and brain chemistry. A medial position was also chosen as lateral voxels can be harder to place due to tissue boundaries. The voxel was placed manually by referring to the individual subject’s T1-weighted (MPRAGE) scan. Specifically, we ensured that the voxel was placed very close to the mid-line of the brain, and as anterior as possible whilst avoiding gyri and cerebrospinal fluid. As such, the voxel was placed both anterior and slightly dorsal to the corpus callosum. Spectra were acquired using a Spin ECho full Intensity-Acquired Localized spectroscopy (SPECIAL; Mlynarik et al., 2006) ^1^H-MRS sequence with water suppression (TR = 3000ms; TE = 8.5ms; Phase cycle Auto; 192 averages from the right prefrontal cortex voxel (Godlewska et al., 2015). Water un-supressed spectra (16 averages) were also acquired. Outer volume suppression slabs were applied 5mm from the edge of each side of the voxel (six slabs in total), both to suppress signals originating outside of the right medial prefrontal voxel of interest, and to minimize motion artefact effects on spectra within the voxel.

**Figure 1.**
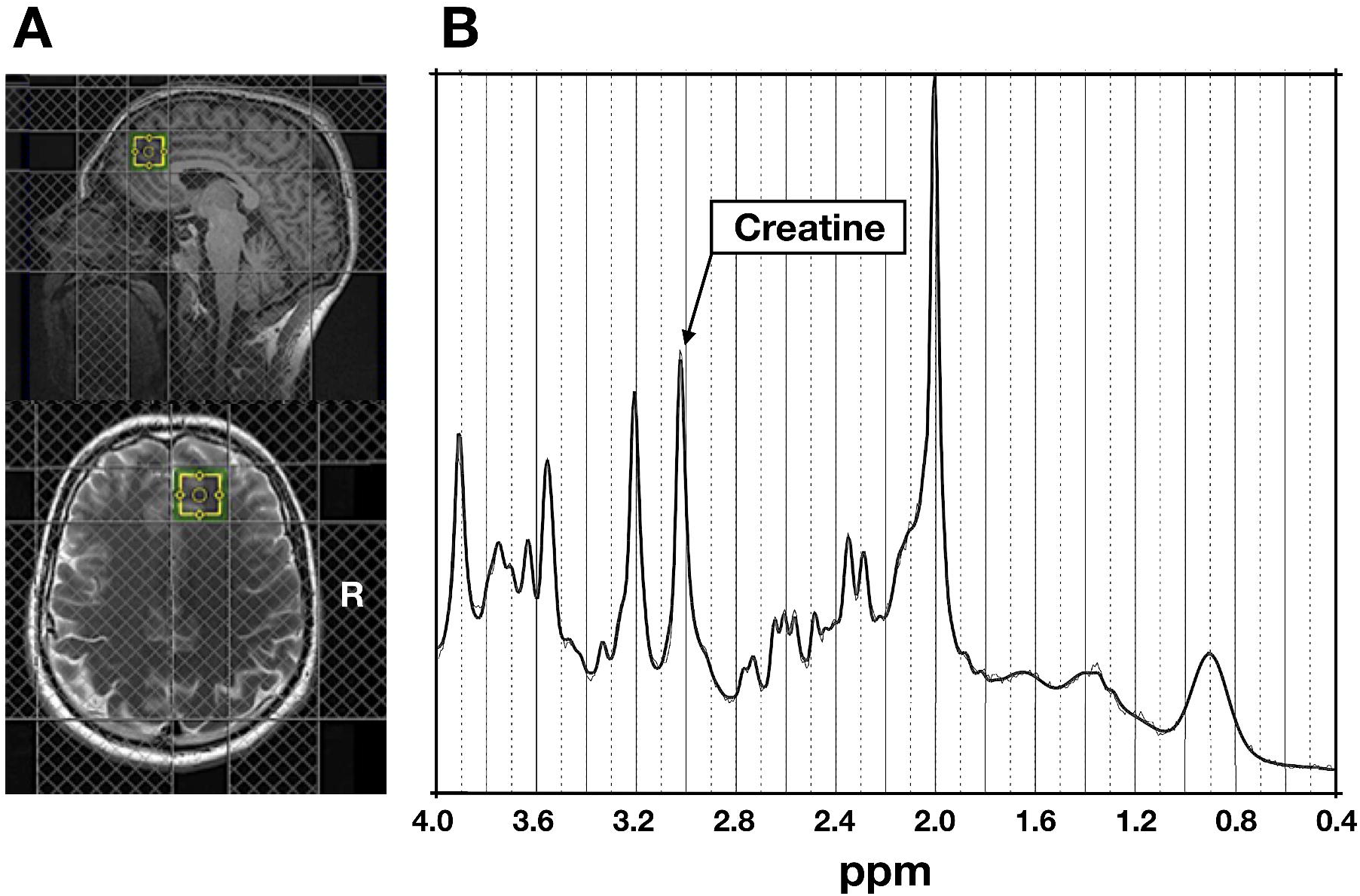
**A:** Typical ^1^H-MRS voxel placement in the medial prefrontal cortex. **B**: Example attained spectrum from the medial prefrontal voxel seen in A.

Spectra were analysed using LCModel 6.3-1L, with a basis set consisting of 19 simulated spectra, as in Morgenroth et al (2019) and Faulkner et al (2020a) (for the full basis set, see Supplementary Materials). This basis set was simulated using FID-A (Simpson et al., 2017) for TE = 8.5ms, magnetic field strength = ∼3T and assuming ideal RF pulses. Line widths and signal-to-noise ratios were estimated as less than 8Hz and greater than 40, respectively (Godlewska et al., 2015). Cramér-Rao lower bounds (Kreis, 2016), line widths and signal-to-noise ratios did not differ as a function of DASS scores (i.e. levels of depression, anxiety or stress) or between study 1 and study 2 (see supplementary materials).

Water referencing and eddy current correction were used to quantify metabolite levels. When quantified in this way, such levels are influenced by cerebral spinal fluid, gray and white matter volumes of the region in which spectra are obtained (Srinivasan et al., 2006), as well as by individual differences in whole-cortical gray matter (Huster et al., 2007). We therefore corrected these metabolite levels for gray and white matter content within the right medial prefrontal voxel using the GABA Analysis Toolkit (Gannet 3.1, http://gabamrs.blogspot.co.uk/), adapted to work with Siemens SPECIAL data. Segmentation was performed using ‘new segment’ in SPM12 (www.fil.ion.ac.uk/spm/software/spm8/). Cerebrospinal fluid, gray and white matter volumes were then accounted for in the expression of creatine using LCModel (Ernst et al., 1993, Gasporovic et al., 2006). For the specific calculation by which metabolites were corrected for using these volumes, see the supplementary materials.

To determine relationships between metabolite concentrations and DASS depression, anxiety or stress scores, an ANOVA was constructed in which DASS depression scores were added as the dependent variable, and age, gender, the average number of cigarettes smoked per day and daily cannabis use were all added as separate factors. For completeness, two exploratory ANOVAs were performed in which the dependent variables were 1) DASS anxiety scores and 2) DASS stress scores, to determine relationships between both anxiety and stress respectively and metabolite concentrations; these two ANOVAs both corrected for age, gender, daily cigarette smoking and daily cannabis use. Because the primary hypothesis pertained *only* to the relationship between depression and prefrontal creatine, and because all other analyses were considered secondary, corrections for multiple comparisons were not applied.

### sMRI Data Acquisition, Pre-processing and Analysis

In both studies, high-resolution structural images were acquired using a T1-weighted magnetization-prepared rapid gradient echo (MPRAGE) sequence. Images were analysed using Computational Anatomy Toolbox 12 (CAT12; http://www.neuro.uni-jena.de/cat) implemented in SPM12 (Wellcome Trust Centre for Neuroimaging; www.fil.ion.ac.uk/spm/software/spm12), as per the standard protocol (see http://www.neuro.uni-jena.de/cat12/CAT12-Manual.pdf); for a detailed description of the sMRI data pre-processing steps, see the supplementary materials.

To determine the relationship between DASS depression scores and whole-brain gray matter volume, group-level analyses were performed by constructing a one-sample general linear model (GLM) in SPM12 that contained each participant’s modulated, normalized, segmented, registered and smoothed gray matter tissue segments, and one explanatory variable (DASS depression scores), along with separate variables for age, gender, daily cannabis use, smoking status (smokers vs non-smokers) and total intracranial volume to control for the influence of these variables. Contrasts were performed to identify regions in which whole-brain gray matter volume A) positively and B) negatively correlated with DASS depression scores. For completeness, two exploratory GLMs were performed; one that included DASS anxiety scores, and one that included DASS stress scores in place of depression scores. A threshold of *p* < 0.05 with FWE correction for multiple comparisons was applied to all contrasts.

To determine relationships between metabolite concentrations and gray matter volume that was associated with depression severity, bivariate correlations were performed; these models included only metabolite concentrations and values from significant clusters that were identified from the above volumetric contrasts.

All ANOVAs and correlational analyses were performed using both frequentist and Bayesian analyses. Frequentist analyses were performed using the Statistical Package for Social Scientists version 26 (SPSS Inc., Chicago, Illinois). Bayesian analyses were performed using JASP (JASP Team (2019), version 0.11.1).

For the frequentist analyses, a significance threshold of alpha = 0.05 (2-tailed) was adopted. For the Bayesian analyses, we adopted the thresholds set out by Jeffreys (1961); for a detailed description of these thresholds, see the supplementary materials.

## Results

### Participant Characteristics

Of the 84 participants across both studies, 46 were male and 38 were female (mean age = 23.42 years, SD = 4.50 years, range = 18 – 37 years). Forty-four participants self-reported tobacco use at least one day per week (mean cigarettes smoked per day by these 44 participants = 5.37, SD = 5.85). Forty-one participants self-reported cannabis use at least once per week (mean number of joints smoked per day by these 41 participants = 0.81, SD = 1.31). Importantly, bivariate correlation analyses revealed that depression scores were not associated with either the number of cigarettes smoked per day (*r* = −0.180, *p* = 0.175, *BF*_*10*_ = 0.203) or daily cannabis use (*r* = −0.231, *p* = 0.835, *BF*_*10*_ = 0.091). A full summary of participant characteristics can be seen in Table 1.

**Table 1.**
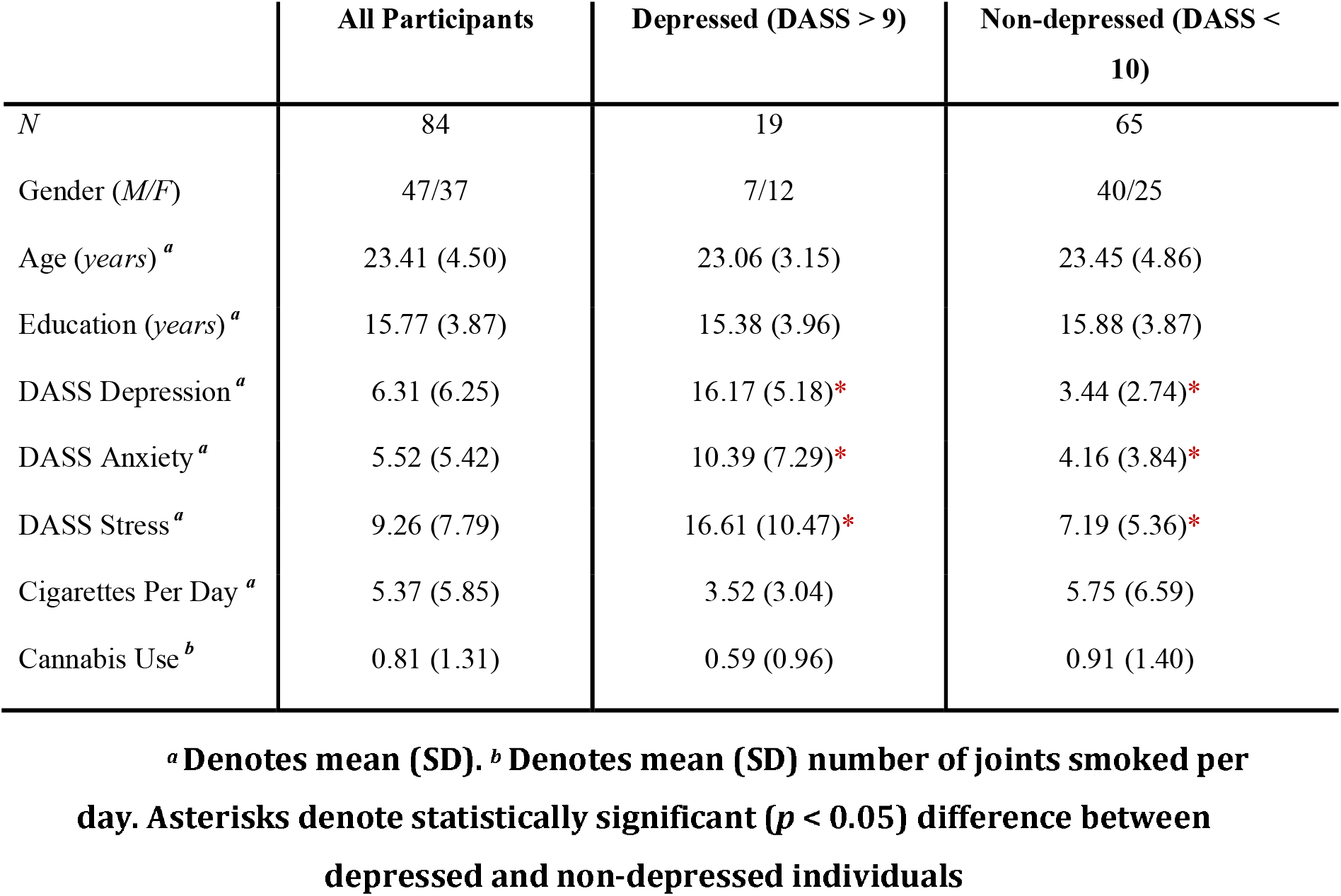
Participant Characteristics.

<sup>a</sup> Denotes mean (SD). <sup>b</sup> Denotes mean (SD) number of joints smoked per day. Asterisks denote statistically significant ( $p < 0.05$ ) difference between depressed and non-depressed individuals
|  | All Participants | Depressed (DASS > 9) | Non-depressed (DASS < 10) |
| --- | --- | --- | --- |
| *N* | 84 | 19 | 65 |
| Gender (*M/F*) | 47/37 | 7/12 | 40/25 |
| Age (*years*)<sup>*a*</sup> | 23.41 (4.50) | 23.06 (3.15) | 23.45 (4.86) |
| Education (*years*)<sup>*a*</sup> | 15.77 (3.87) | 15.38 (3.96) | 15.88 (3.87) |
| DASS Depression<sup>*a*</sup> | 6.31 (6.25) | 16.17 (5.18)* | 3.44 (2.74)* |
| DASS Anxiety<sup>*a*</sup> | 5.52 (5.42) | 10.39 (7.29)* | 4.16 (3.84)* |
| DASS Stress<sup>*a*</sup> | 9.26 (7.79) | 16.61 (10.47)* | 7.19 (5.36)* |
| Cigarettes Per Day<sup>*a*</sup> | 5.37 (5.85) | 3.52 (3.04) | 5.75 (6.59) |
| Cannabis Use<sup>*b*</sup> | 0.81 (1.31) | 0.59 (0.96) | 0.91 (1.40) |

### Depression Scores

The mean self-reported score on the depression subscale of the DASS was 6.31 (SD = 6.25, range = 0 to 24). Specifically, 65 participants scored between 0-9 and so were classified as ‘non-depressed’ (mean of these 65 participants = 3.28, SD = 2.72), while the remaining 19 participants scored 10+ and were thus classified as ‘depressed’ (mean of these 19 participants = 18.47, SD = 8.25); of these, 8 scored between 10-13 and were defined as ‘mildly depressed’ (mean = 10.33, SD = 0.71), 5 scored between 14-20 and were defined as ‘moderately depressed’ (mean = 17.00, SD = 2.22) and 6 scored between 21-27 and were defined as ‘severely depressed’ (mean = 22.00, SD = 1.20). The skewness and kurtosis values for depression scores were slightly outside the acceptable range, and so a square-root transformation was performed on this variable to render the data normally-distributed.

An ANOVA that contained DASS depression scores as the dependent variable and both age and gender as separate factors revealed that depression scores were not influenced by age (*F*(1,81) = 0.585, *p* = 0.447, *BF*_*10*_ = 0.666). However, this ANOVA did reveal that female participants reported higher DASS depression scores than male participants (*F*(1,81) = 6.446, *p* = 0.013, *BF*_*10*_ = 1.431; *see Figure S1*).

### Relationship Between DASS Depression Scores and Prefrontal Creatine and Other Metabolites

An ANOVA that controlled for age, gender, daily cigarette and cannabis use, revealed that there was an association between DASS depression scores and concentrations of creatine in the prefrontal cortex (*F*(1,76) = 6.005, *p* = 0.017; *BF*_*10*_ = 3.834; *see Figure 2*); specifically, participants who self-reported the highest depression scores exhibited the lowest creatine concentrations in the medial prefrontal cortex. There were no significant relationships between DASS depression scores and concentrations of any of the remaining metabolites (see supplementary materials). For the effects of age, gender, cannabis and tobacco use on metabolite concentrations, see the supplementary materials.

**Figure 2.**
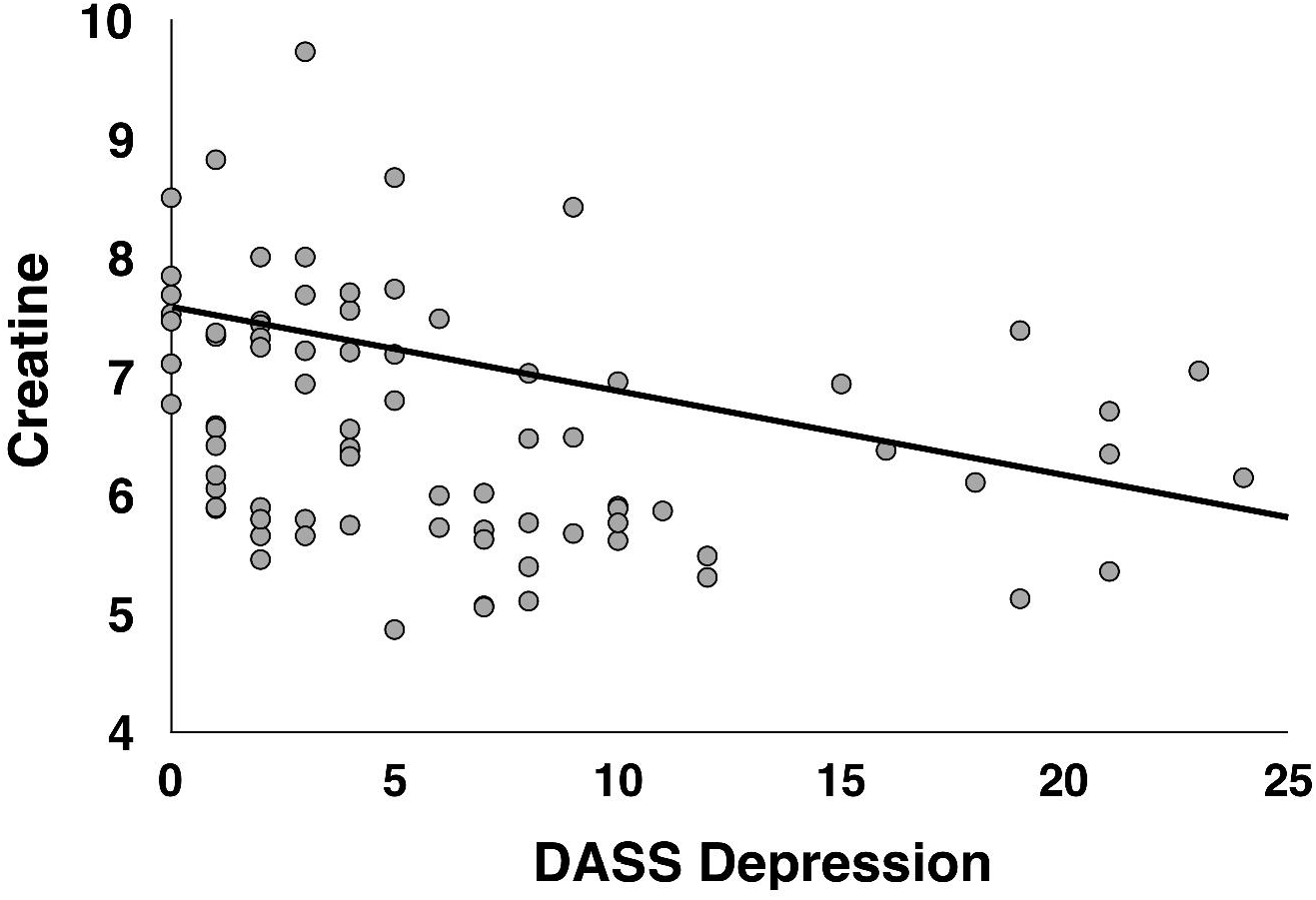
Relationship between DASS depression scores and Creatine (corrected for gray matter) in the prefrontal voxel depicted in Figure 1A.

### Relationship Between Depression Scores and Gray Matter Volume

When controlling for age, gender, daily cigarette use, daily cannabis use and total intracranial volume, there was a significant negative association between DASS depression scores and gray matter volume in a cluster of 524 voxels in the right medial superior frontal gyrus; the local maxima for this cluster was *x* = 2, *y* = 58, *z* = 16 (see Figure 3).

**Figure 3.**
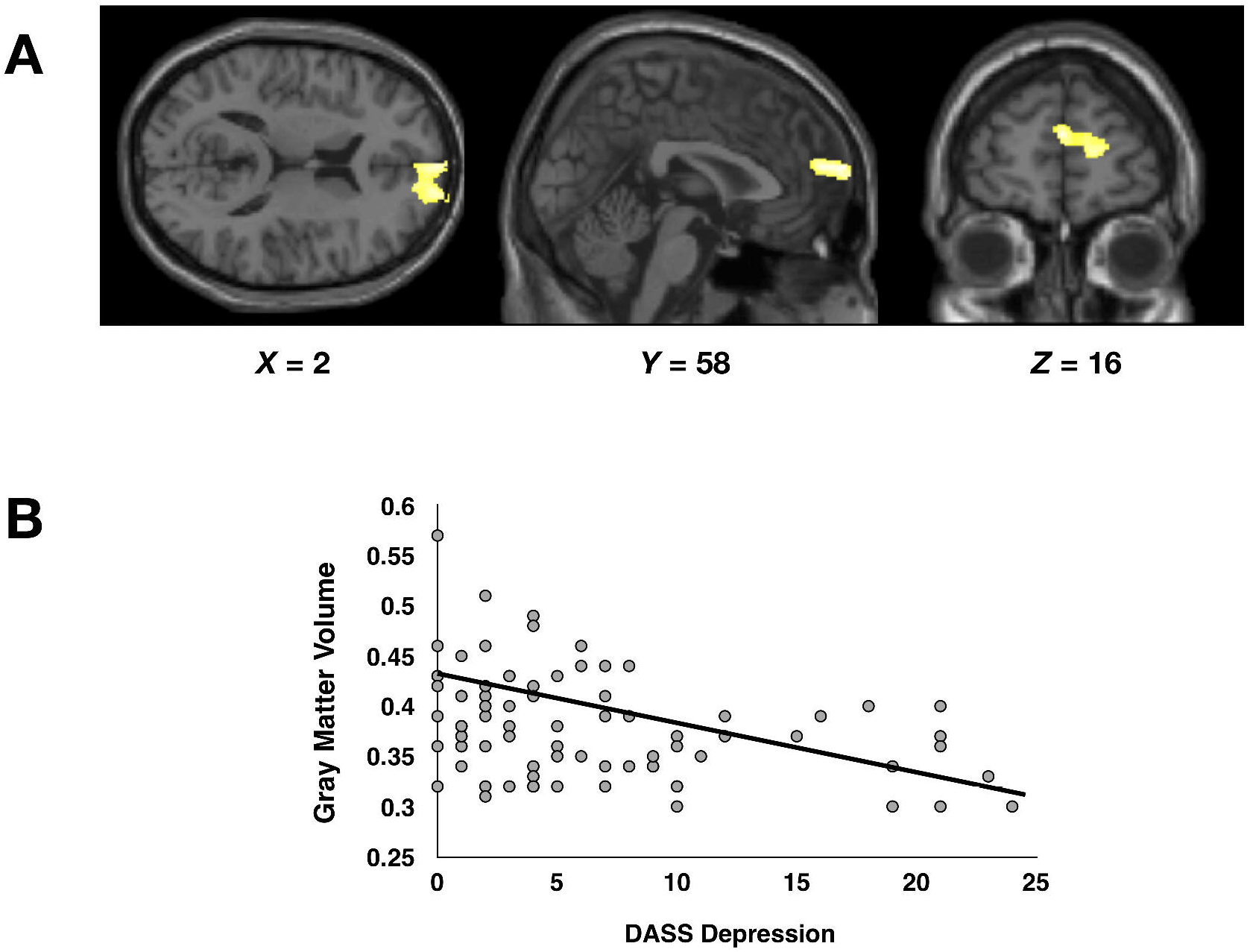
**A**: Results of the whole-group F-test that examined brain regions in which whole-brain gray matter was associated with DASS depression scores. **B:** Scatterplot depicting the relationship between DASS depression scores and gray matter volume within the cluster of voxels depicted in A.

**Figure 4.**
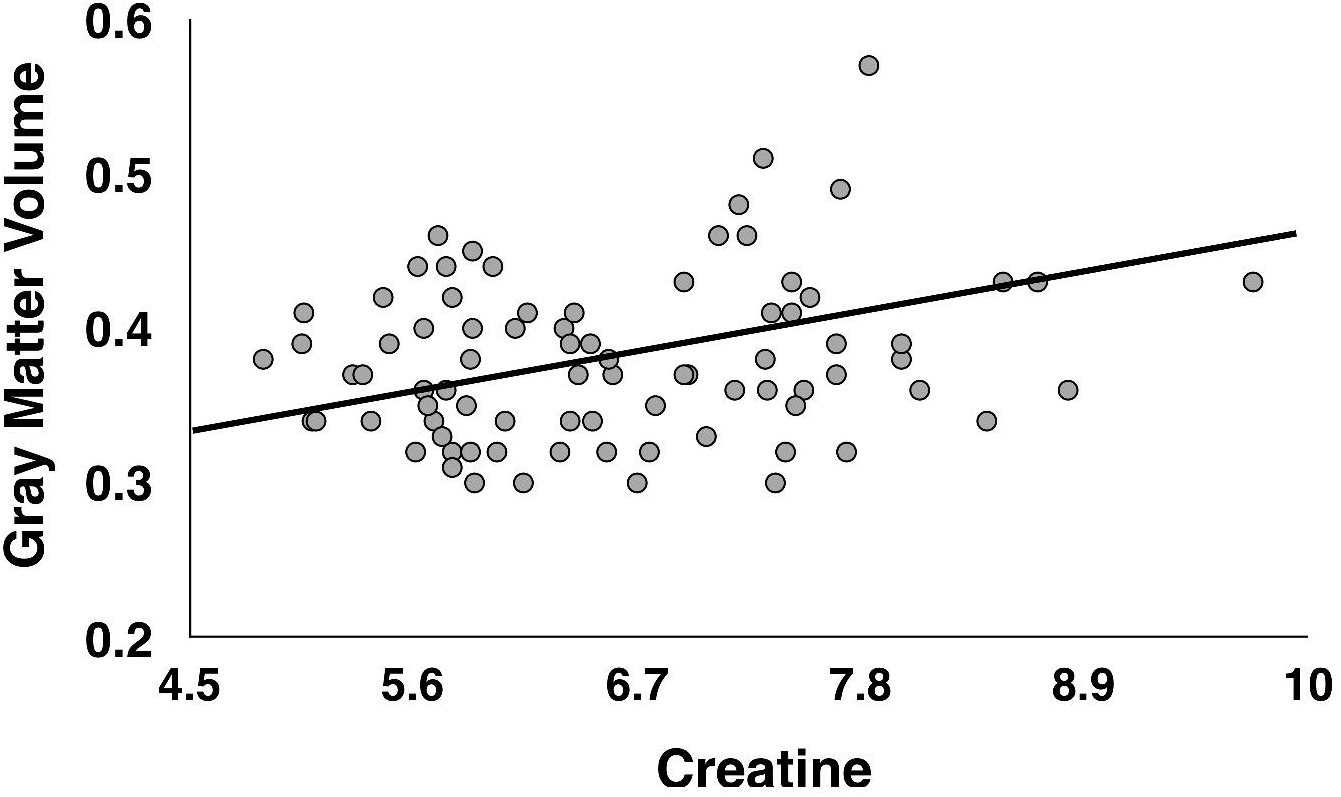
Correlation between concentrations of Creatine in the voxel shown in Figure 1A, and gray matter volume in the cluster of voxels shown in Figure 3A in all participants.

### Relationship Between Prefrontal Creatine and Gray Matter Volume

A bivariate correlation revealed a significant positive correlation between prefrontal creatine concentrations and values extracted from the cluster of 524 voxels depicted in Figure 3A (values averaged across all voxels in the cluster) (*r* = 0.225, *p* = 0.043, *BF* = 3.128). Finally, none of the remaining metabolite concentrations correlated with values extracted from the cluster of voxels depicted in Figure 3A (all *p*s > 0.187, all *BF*s < 0.919).

### Anxiety and Stress Scores

Partial correlation analyses that controlled for age, gender, daily cigarette use and daily cannabis use revealed that neither DASS anxiety scores nor DASS stress scores correlated with concentrations of creatine or any of the remaining metabolites (all *p*s > 0.163, all *BF*_*10*_ < 0.802; see supplementary materials). Finally, there were no significant associations between whole-brain gray matter volume and scores on the DASS anxiety or stress subscales.

## Discussion

This is the first study to examine the relationship of both prefrontal creatine and gray matter volume with depression/low mood in a group of individuals who self-reported a wide range of depression severities. Our results suggest that individuals who experience sub clinical depressive symptoms and low mood have lower concentrations of creatine and lower gray matter volume in the prefrontal cortex. These findings extend those of previous research, by indicating that depression, when considered as a continuum that ranges from no/sub-clinical depression to severe depression, is associated with low creatine and gray matter volume in the prefrontal cortex.

That greater depression severity was related to lower concentrations of prefrontal creatine was expected on the basis of findings from one study, which revealed a weak negative relationship between depression and frontal creatine (measured using 31-phosphorous MRS) in a small sample of adolescent depressed participants (Kondo et al., 2016). Interestingly, the relationship between brain creatine and magnitude of depression may be specific to the prefrontal lobes, as previous studies indicate there to be no relationship between depression and concentrations of creatine in the thalamus (Mirza et al., 2005) or anterior cingulate cortex (Auer et al., 2000).

The present findings could partly explain why increasing creatine levels via administration of creatine supplements has been shown to have potential in alleviating depressive symptoms in depressed patients (Kious et al., 2019; Kondo et al., 2011; Lyoo et al., 2012; Nemets and Levine, 2013; Roitman et al., 2007). However, while Kondo et al (2016) examined the effects of daily administration of 2mg, 4mg or 10mg creatine on both depression and prefrontal creatine, they did so in only seven, eight and seven participants respectively, meaning that the authors may have been underpowered to determine whether baseline prefrontal creatine concentrations can predict the observed treatment-induced improvements in mood. Future studies may wish to use magnetic resonance spectroscopy to determine whether pre-treatment concentrations of creatine can predict such a response, and whether this effect is indeed specific to the prefrontal lobes, in a larger sample size than used by those authors.

Currently, the mechanisms by which depression and low mood are associated with low brain creatine are poorly understood. Creatine has an important role in the regulation of many neurological functions including sodium and calcium transport, and the synthesis, uptake and release of neurotransmitters (Allen, 2012). Importantly, impairments in all of these functions are considered to promote depression (Assis et al., 2009; Kious et al., 2019). Following ingestion or biosynthesis, creatine is transferred from blood plasma to the brain via chloride-dependent creatine transporters (CRTs); this may be one way by which daily administration of 20mg creatine monohydrate can increase total creatine in cortical and subcortical gray and white matter (see Allen (2012) for a review). Further, many of the brain regions that express CRTs, including the prefrontal cortex, are compromised in depression (e.g. Allen, 2012). Furthermore, administration of creatine can increase brain-derived neurotrophic factor (BDNF) in the hippocampus, which has itself been shown to have antidepressant effects (Pazini et al., 2016). However, these cellular mechanisms cannot be observed using ^1^H-MRS as this technique does not distinguish between intracellular and extracellular concentrations of creatine, nor does it allow for observation of creatine function at the level of the transporter. To truly determine the influence of creatine function (and indeed of creatine supplements) on depression, future studies could utilize other neuroimaging techniques, such as Positron Emission Tomography, alongside ^1^H-MRS and structural MRI, to better examine prefrontal creatine function and affect at many stages throughout an individual’s lifespan, and to determine whether creatine supplements can alter mood via actions in the prefrontal cortex.

The present findings indicate that levels of depression, especially when quantified as in this study may be related to concentrations of creatine in the prefrontal voxel observed in Figure 1A, but not with other neurotransmitters in this specific brain region. Interestingly, while previous research (e.g. Auer et al., 2000) has used ^1^H-MRS to indicate that clinical depression is associated with low concentrations of glutamate in other brain regions such as the anterior cingulate cortex (which is considered to be adjacent to the prefrontal cortex), studies that have examined the relationship between depression and glutamate specifically in the prefrontal cortex have often corrected glutamate for concentrations of creatine, and have only examined the relationship of such concentrations with clinical depression (Hasler et al., 2007). As such, our results indicate that when examining the relationship between levels of depression that range from almost zero depression to high depression (which may be considered socially-representative) and metabolite concentrations in the prefrontal cortex, the former are associated with concentrations of creatine, rather than glutamate. However, future work is needed to determine the replicability of this particular finding. Furthermore, our results also indicate that low prefrontal creatine and gray matter volume are specifically related to depression, as the DASS allows for identification of various aspects of emotional disturbance, and no meaningful or significant relationships between our brain measures and scores on the DASS anxiety or stress subscales were found.

The results of our VBM analyses replicate findings from previous studies (e.g. Bora et al., 2012; Lai et al., 2013), by indicating that higher levels of depression are associated with lower gray matter volume in the right superior medial frontal gyrus. As behavioural and cognitive data were not available from a sufficient sample of participants, our data cannot determine the functional relevance of reductions in gray matter in this brain region. However, low volume within the superior medial frontal gyrus may be associated with known depression-related cognitive deficits. For example, low gray matter volume and hypoactivation within this region are related to high levels of rumination (Johnson et al., 2009; Schiller et al., 2013) and an inability to inhibit a prepotent response (Li et al., 2010), both of which are associated with depressive symptoms (e.g. Kaiser et al., 2003 Nolen-Hoeskema, Parker and Larson, 1994). Future studies could therefore aim to determine whether therapies that act upon/prevent damage to this brain region can aid in the treatment of depression/low mood.

This study has several limitations. Firstly, participants were typically young adults, and so our findings may not be generalizable to the entire public. Secondly, only 19 participants were classified as ‘depressed’ by the DASS because they scored greater than 9 on the depression subscale, meaning that we may have lacked sufficient statistical power to detect small effects. Similarly, a larger sample size would have provided greater statistical power to detect small changes in brain metabolites such as Taurine that have less-dominant peaks and thus lower signal-to-noise. Thirdly, a structured clinical interview (such as the Mini-International Psychiatric Interview (Sheehan et al., 1998), for example) was not administered, meaning that detailed information regarding clinical symptoms of depression could not be determined. Fourthly, our cross-sectional design did not allow us to make predictions regarding the causal effects of brain metabolites on depression; future studies may therefore wish to employ a longitudinal design. Fifthly, some participants self-reported light-to-moderate use of cannabis, and moderate use of tobacco cigarettes; while our analyses indicated cannabis use did not influence brain creatine or gray matter volume, there is a possibility that it had a small, yet undetectable effect. In addition, because our ^1^H-MRS sequence only allowed for the quantification of metabolite concentrations in one voxel (see Figure 1), we were not able to determine whether level of depression is related to creatine (or indeed any other metabolite) in brain regions outside of the medial prefrontal cortex. Finally, cognitive measures were not obtained from a sufficient sample of participants, meaning that we could not determine whether low creatine is associated with depression-related cognitive deficits.

In summary, this study provides evidence in a representative group of individuals who exhibit a range of depression levels, that prefrontal creatine and gray matter volume are negatively related to low mood/depression. While future research is needed to achieve a more complete understanding of this relationship, these results support the findings of research which indicate that increasing concentrations of prefrontal creatine via creatine supplements may be able to improve an individual’s mood and/or alleviate depression.

## Data Availability

For availability of this data, please contact Paul Faulkner.

